# Assessing attitude, self-efficacy, and perceived risk toward seasonal influenza vaccination among primary care physicians in Qatar: A cross-sectional study

**DOI:** 10.1101/2025.04.11.25325654

**Authors:** Kamran Aziz, Mansoura Salem Ismail, Marwa Bibars, Nagah Selim, Ayah Mohamed, Ahmed Sameer Alnuaimi, Muna Mehdar AlSaadi

## Abstract

This cross-sectional study examined influenza-related attitude, self-efficacy and perceived risk among 190 primary health care physicians. Participants were predominantly male (over 50%), aged ≥45 years (55.7%), with one-third reporting chronic diseases. Most (96.3%) had prior influenza vaccination, though only 73.3% were vaccinated in the last season. While 87.9% endorsed vaccination for healthcare workers to ensure service continuity, only 37.4% supported mandatory policies. Notably, 46% believed healthcare professionals do not pose influenza transmission risks to patients. Self-efficacy for vaccination was strongly tied to time availability (73.7% agreement) and institutional vaccine provision (78.9%), with social support from colleagues (79.5%) and relatives (68.9%) further influencing adherence. Male physicians (87.5%) and those with ≥5 prior vaccine doses (88.6%) or recent vaccination (87.3%) reported higher self-efficacy, though chronic disease history showed no significant association. Risk perception disparities emerged: 94.2% acknowledged elevated occupational risk during epidemics, yet only 62.1% perceived personal risk. Similarly, 86.3% viewed influenza as dangerous for patients versus 64.2% for themselves. Higher perceived risk scores correlated with chronic disease history (84.5), prior vaccination (81.1), recent vaccination (82.8), and ≥5 vaccine doses (85.0). Information sources prioritized official health agencies (Ministry of Public Health: 59.5%; WHO/CDC: 56.3%), while traditional media were least utilized (7.9– 21.1%). These findings highlight gaps between professional risk acknowledgment and personal risk mitigation, underscoring the need for targeted strategies to address vaccine hesitancy, improve access, and align perceptions with evidence-based practices in healthcare settings.

## 2. Introduction

Seasonal influenza is considered a significant public health concern that leads to sizable morbidity, mortality, and healthcare costs worldwide. It is projected that there are about one billion cases of influenza worldwide per year, of which 3–5 million are severe cases and nearby 0.65 million lead to influenza-related respiratory death [1,2]. Health workers (HWs) are at increased risk of influenza virus infection, which may vary depending on occupation or setting [3]. Infected HWs could increase the risk of nosocomial infection and community spread [4,5]. The seasonal influenza vaccine is one of the most effective preventive measures to reduce the impact of influenza infections. The World Health Organization (WHO) considers HWs to be a priority group for influenza vaccination and reinforces this position by supporting countries to develop and implement national, seasonal immunization policies for HWs, but vaccination rates among healthcare workers, remain suboptimal. Primary care physicians (PCPs) play a critical role in influencing vaccination decisions, both for their patients and for themselves. However, the success of vaccination programs often depends on the attitudes, perceived risks, and self-efficacy of healthcare providers, including PCPs. [6,9]. PCPs’ attitudes toward the seasonal influenza vaccine can influence their recommendation and administration of the vaccine to their patients. Studies suggest that healthcare professionals’ personal attitudes and beliefs about vaccines are significant predictors of their vaccination practices. Positive attitudes toward vaccination can increase vaccination rates. PCPs’ perception of the risks associated with influenza—also plays a crucial role in vaccination uptake [10,12]. Research has shown that healthcare workers who perceive themselves at higher risk for contracting or transmitting influenza are more likely to get vaccinated. Self-efficacy, or the belief in one’s ability to execute a behavior successfully, is another key factor influencing vaccination practices. PCPs with high self-efficacy regarding the influenza vaccine are more likely to recommend it to patients and receive it themselves. Self-efficacy can be shaped by various factors, including training, knowledge of the vaccine’s benefits, and previous positive experiences with vaccination. Enhancing PCPs’ self-efficacy through education and training can lead to higher vaccine uptake rates [13,15]. Understanding PCPs’ attitudes, perceived risks, and self-efficacy regarding the seasonal influenza vaccine is vital for improving vaccination coverage. Effective vaccination campaigns must address the factors that influence healthcare providers’ vaccine behaviors, as they serve as trusted sources of information for patients. By focusing on improving attitudes and self-efficacy, public health initiatives can enhance influenza vaccination rates, ultimately reducing the burden of influenza-related morbidity and mortality.

## 3. Method of study

An online survey of all primary healthcare physicians working in primary healthcare corporation in Qatar was conducted from October 11, 2021, to October 31, 2021.

A validated questionnaire was obtained from Asma et al. [16] study about background characteristics such as age, gender, working experience in primary care, and history of influenza vaccination… The second section included attitude, self-efficacy, and perceived risk regarding influenza vaccination, which were assessed using five-point Likert questions. Responses were expressed as follows: 1 means totally agree, 2 means I agree, 3 means neutral, 4 means I disagree, and 5 means I totally disagree. The percentage reported was for those who answered as agree or strongly agree and each positive item was awarded a score of one point. The total score was calculated by summing the scores of positive items and multiplying it by 100/maximum count of items.

### 3.1 Data analysis

The collected data was analyzed using SPSS version 25 and the quality of the data was checked before proceeding to the analysis. Quantitative data was summarized in the form of mean and SD while qualitative data was summarized in the form of frequency tables and charts. The Bivariate analysis was then carried out using the independent t-test for two groups and ANOVA for more than two groups to investigate the association between the dependent variables and the background characteristics of physicians. Effect size is assessed by Cohen’s d for differences in the mean between two groups, while Eta squared test was used for the effect of a factor with more than two groups. The level of statistical significance was set at a p-value of less than or equal to 0.05.

### 3.2 Ethics statements

This was an anonymous, online-based survey utilizing implied informed consent. Participants were informed that by submitting the completed questionnaire, they were providing their consent to participate. A survey link was distributed to all physicians, accompanied by a statement indicating that clicking the link signified voluntary consent to take part in the study. The questionnaire included a cover page outlining the study’s aims and objectives, emphasizing that participation was entirely voluntary and that no identifiable information would be collected. All anonymized responses were received directly by the researcher.

### 3.3 Ethics approval

The Institutional Review Board at Primary Health Care Corporation in Qatar approved this study (Reference number PHCC/DCR/ 2020/06/047

## 4. Results

The overall response rate in this study was 42%, 106 (55.7%) were 45 years old and above, and more than half of the participants were males. About one-third reported a history of Chronic diseases [Table 1].

**Table 1:** background characteristics of physicians working in Primary healthcare centers in Qatar.

| Characteristic | N | % |
| --- | --- | --- |
| Age |  |  |
| ≤44 | 84 | 44.2 |
| ≥ 45 | 106 | 55.7 |
| Female | 78 | 41.1 |
| Male | 112 | 58.9 |
| Duration of PHCC work experience in years |  |  |
| <8 | 134 | 70.5 |
| ≥8 | 56 | 29.5 |
| History of chronic diseases |  |  |
| No | 129 | 67.9 |
| Yes | 61 | 32.1 |
| Total | 190 | 100 |

Most participants (96.3%) had been vaccinated against influenza in the past, while 73.3% of the physicians had received an influenza vaccine in the last season [Table 2].

**Table 2:** History of Influenza vaccine uptake among primary care healthcare in Qatar.

|  |  |  |
| --- | --- | --- |
| Ever taken influenza vaccination |  |  |
| No | 7 | 3.7 |
| Yes | 183 | 96.3 |
| influenza vaccination last season |  |  |
| No | 50 | 26.3 |
| Yes | 140 | 73.7 |
| Doses of seasonal influenza vaccine |  |  |
| Never | 7 | 3.7 |
| (1-2) | 25 | 13.2 |
| (3-4) | 49 | 25.8 |
| ≥5 | 87 | 45.8 |
| Don't remember | 22 | 11.6 |
| Total | 190 | 100.0 |

Of 190 doctors, the majority (87.9%) believe that health professionals should be vaccinated for the continuity of health services. Despite this, only 37.4% believe that the influenza vaccine should be mandatory for healthcare workers. Notably, nearly 46% of the respondents feel confident that health professionals do not pose a risk of spreading the influenza virus to their patients [Table 3].

**Table 3:** Attitude of primary care physicians in Qatar towards Influenza vaccination (*N* =190).

|  | n | % | 95%confidence interval |
| --- | --- | --- | --- |
| I feel that health professionals are not spreading the disease to their patients | 87 | 45.8 | (38.8-52.9) |
| I believe that health professionals should be vaccinated for the continuity of health services | 167 | 87.9 | (82.7-92) |
| Influenza vaccine should be mandatory for health professionals | 71 | 37.4 | (30.7-44.4) |

When asked about their self-efficacy in successfully getting vaccinated, Time and availability were the two most important factors. 73.7% agreed that they would be vaccinated if they had enough time while 78.9% expressed willingness to be vaccinated if the institute provided the vaccine. Perceived social support for influenza vaccination from relatives and colleagues was frequently reported with individual self-efficacy (68.9% & 79.5% respectively) [Table 4].

**Table 4:** Self-efficacy of primary care physicians towards Influenza vaccination (*N* =190).

|  | n | % | 95%confidence interval |
| --- | --- | --- | --- |
| My relatives believe that my vaccination is important | 131 | 68.9 | (62.1 - 75.2) |
| My institute recommends being vaccinated | 185 | 97.4 | (94.3 - 99) |
| My colleagues believe that my vaccination is important | 151 | 79.5 | (73.3 - 84.7) |
| The Ministry of Health recommends vaccination of health professionals | 187 | 98.4 | (95.8 - 99.6) |
| The health authorities I know recommend vaccination | 182 | 95.8 | (92.2 - 98) |
| I would be vaccinated every year if I have enough time | 140 | 73.7 | (67.1 - 79.6) |
| I would be vaccinated if someone reminds me | 126 | 66.3 | (59.4 - 72.7) |
| I would be vaccinated every year if the vaccine is provided in my institute | 150 | 78.9 | (72.7 - 84.3) |
| I would be vaccinated every year if I am rewarded | 64 | 33.7 | (27.3 - 40.6) |
| I would be vaccinated for influenza every year if sufficient knowledge was given | 125 | 65.8 | (58.8 - 72.3) |

Significant positive associations with self-efficacy mean scores were observed among primary care physicians who were male (87.5%) and those who had received five or more influenza vaccinations (88.6%), particularly in the most recent season (87.3%). In contrast, there was no statistically significant relationship between a history of chronic diseases and self-efficacy scores [Table 5].

**Table 5:**
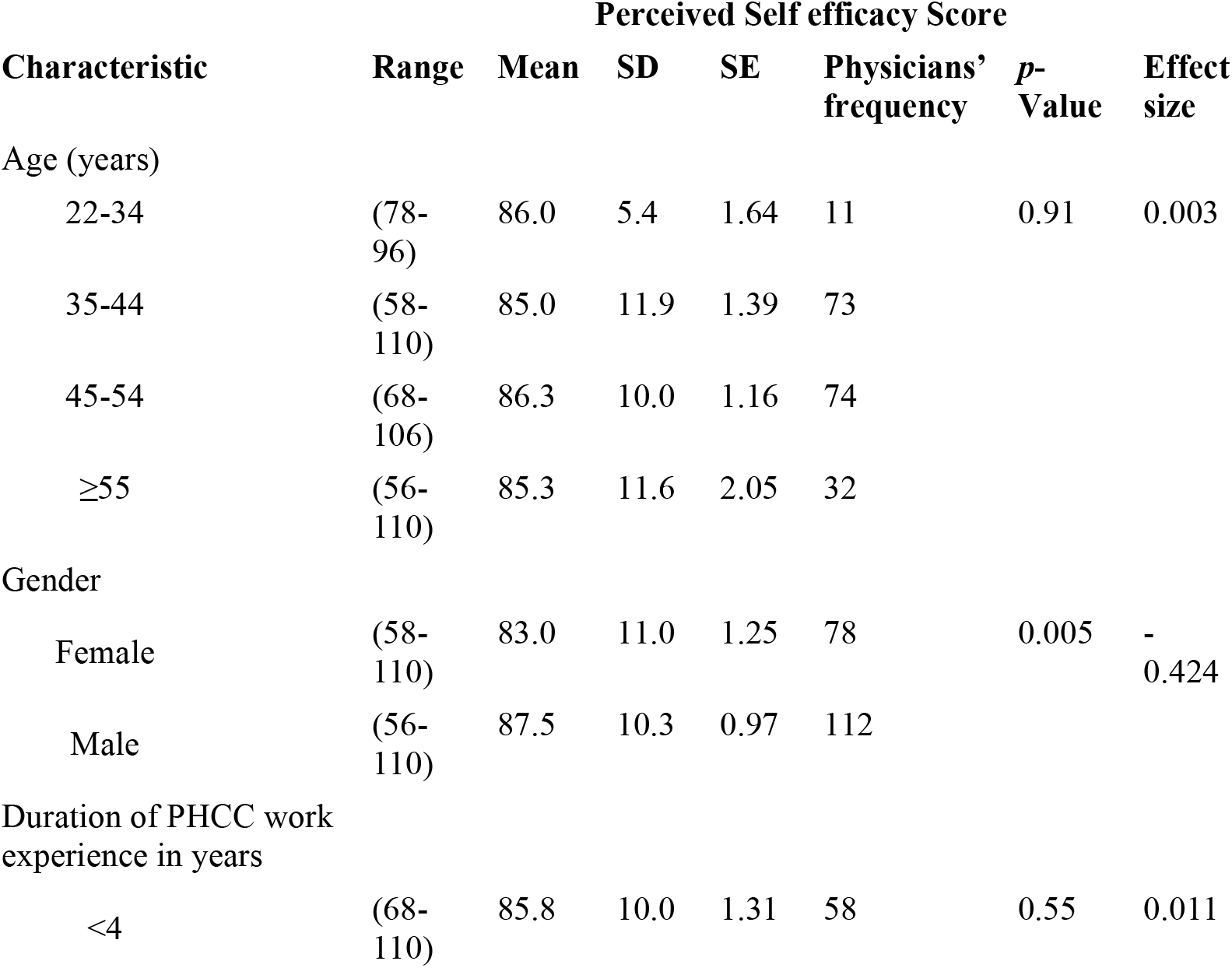
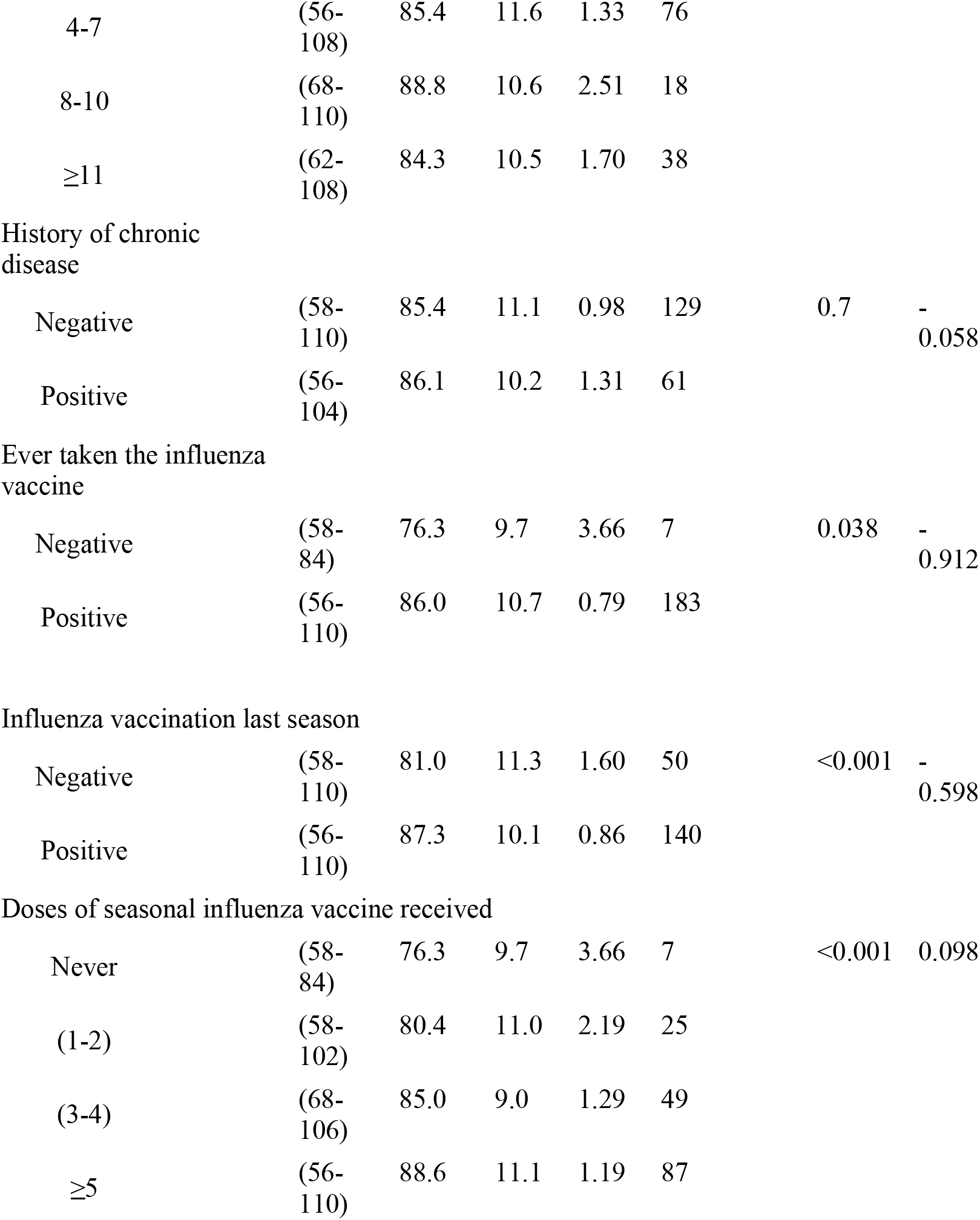
Relationship between sociodemographics of primary care physicians and perceived self-efficacy score in Qatar 2021 (*N* =190).

In assessing risk perception, a difference was reported between physicians’ acknowledgment of risk and their personal risk perception. While 94.2% agreed healthcare professionals face the greatest influenza risk during epidemics, only 62.1% believed they were at high risk. This was reflected in perceived severity; 86.3% considered influenza dangerous for patients, versus only 64.2% for themselves [Table 6].

**Table 6:** Seasonal influenza vaccine perceived risk and severity of risk among primary care physicians (*N* =190).

|  | n | % | 95%confidence interval |
| --- | --- | --- | --- |
| Perceived risk |  |  |  |
| I'm at high risk for influenza | 118 | 62.1 | (55.1 - 68.8) |
| I can spread infection to my patients even if I am asymptomatic | 153 | 80.5 | (74.5 - 85.7) |
| Health professionals are under the highest risk in case of an epidemic | 179 | 94.2 | (90.2 - 96.9) |
| I can spread infection to my family even if I am asymptomatic | 170 | 89.5 | (84.5 - 93.2) |
| Perceived severity of risk |  |  |  |
| Influenza is dangerous for me | 122 | 64.2 | (57.2 - 70.8) |
| Influenza is dangerous for my patients | 164 | 86.3 | (80.9 - 90.6) |
| Influenza is dangerous for my family | 148 | 77.9 | (71.6 - 83.3) |

The analysis revealed a statistically significant association between physicians’ perceived risk mean scores and select background characteristics. Notably, higher mean scores were observed among those with a history of chronic diseases (84.5), prior influenza vaccination (81.1), influenza vaccination in the last season (82.8), and having received five or more seasonal influenza vaccine doses (85.0) [Table 7].

**Table 7:** Relationship between sociodemographic of primary care physicians and perceived risk score in Qatar 2021 (*N* =190).

| Characteristic | Range | Mean | SD | SE | Physicians' frequency | p-Value | Effect size |
| --- | --- | --- | --- | --- | --- | --- | --- |
| Age (years) |  |  |  |  |  | 0.18[NS] | 0.026 |
| 22-34 | (40 - 100) | 75.6 | 15.3 | 4.61 | 11 |  |  |
| 35-44 | (49 - 100) | 79.1 | 12.8 | 1.5 | 73 |  |  |
| 45-54 | (46 - 100) | 82.5 | 12.4 | 1.44 | 74 |  |  |
| ≥55 | (60 - 100) | 82.1 | 11.5 | 2.03 | 32 |  |  |
| Gender |  |  |  |  |  | 0.24[NS] | -0.176 |
| Female | (40 - 100) | 79.4 | 13.2 | 1.49 | 78 |  |  |
| Male | (46 - 100) | 81.6 | 12.2 | 1.16 | 112 |  |  |
| Duration of PHCC<br>work experience in<br>years |  |  |  |  |  | 0.45[NS] | 0.014 |
| <4 | (40 - 100) | 78.9 | 13.1 | 1.73 | 58 |  |  |
| 4-7 | (46 - 100) | 80.8 | 12.7 | 1.46 | 76 |  |  |
| 8-10 | (57 - 100) | 84 | 12.5 | 2.94 | 18 |  |  |
| ≥11 | (60 - 100) | 81.8 | 11.7 | 1.9 | 38 |  |  |
| Having chronic disease |  |  |  |  |  | 0.70[NS] | -0.058 |
| Negative | (40 - 100) | 78.9 | 13.2 | 1.16 | 129 |  |  |
| Positive | (63 - 100) | 84.5 | 10.5 | 1.35 | 61 |  |  |
| Positive history of<br>influenza vaccination |  |  |  |  |  | <0.001 | -0.912 |
| Negative | (63 - 77) | 71 | 6.5 | 2.45 | 7 |  |  |
| Positive | (40 - 100) | 81.1 | 12.7 | 0.94 | 183 |  |  |
| Positive history of influenza<br>vaccination last season |  |  |  |  |  | <0.001 | -0.640 |
| Negative | (40 - 100) | 75 | 13.5 | 1.91 | 50 |  |  |
| Positive | (46 - 100) | 82.8 | 11.7 | 0.99 | 140 |  |  |
| Count of seasonal influenza vaccine doses received |  |  |  |  |  | <0.001 | 0.134 |
| Never | (63 - 77) | 71 | 6.5 | 2.45 | 7 |  |  |
| (1-2) | (40 - 94) | 72.5 | 14.4 | 2.88 | 25 |  |  |
| (3-4) | (51 - 100) | 79.2 | 12.7 | 1.81 | 49 |  |  |
| ≥5 | (51 - 100) | 85 | 10.7 | 1.15 | 87 |  |  |

Physicians primarily rely on official websites from the Ministry of Public Health (59.50%), WHO, and CDC (56.30%) for influenza vaccine updates. Conversely, newspapers (21.10%), TV, and flu-specific platforms (7.90%) are the least consulted sources [Fig. 1].

**Fig 1:**
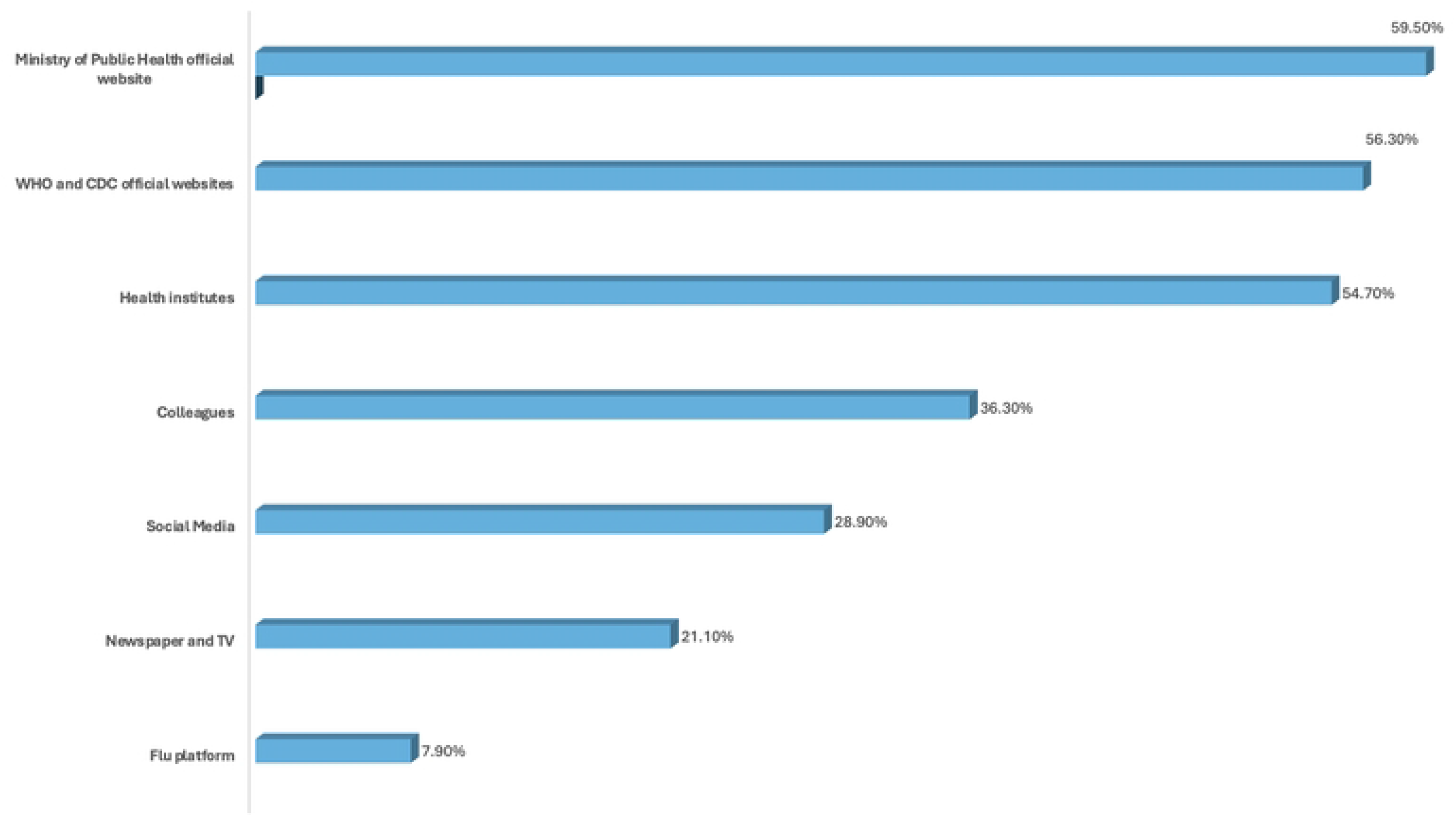
Bar chart for the source of information.

## Discussion

The study focused on primary care physicians’ attitudes, self-efficacy, and perceived risk regarding accepting the seasonal influenza vaccine.

The study revealed that most participants had received an influenza vaccination in the past, while two-thirds of physicians had been vaccinated during the last season. This is per a study done in the US, where family physicians’ vaccination rate was up to 87%, attributed to workplace policies and free access on-site for all staff. [17].

Most physicians believe that healthcare professionals should be vaccinated to ensure the continuity of health services. Studies have shown that physicians know their occupational obligations towards vaccination [18,19].

According to Polan et al. when healthcare professionals are vaccinated against sessional influenza, they increase the ability of the healthcare system to respond effectively to influenza epidemics [20].

About one-third of the physicians thought the influenza vaccine should be mandatory for healthcare professionals. According to Winston et al. mandatory vaccination increases vaccination rates but healthcare professionals negatively perceive it [21].

Approximately half of the respondents felt that health professionals do not contribute to the spread of the influenza virus to their patients. However, some studies suggest that the disease is often asymptomatic, and healthcare professionals can spread the influenza virus to their patients and families [22-24].

The availability of the influenza vaccine and the time needed to get vaccinated were crucial factors influencing individuals’ self-efficacy regarding vaccine uptake. Approximately two-thirds of participants indicated that they would be willing to get vaccinated if they had enough time and access to the vaccine at their institution. AlMarzooqi et al. reported that one of the frequently cited reasons for not taking influenza vaccine was lack of time to take the vaccine (28.9%). The majority (85.1%) of healthcare professionals received the vaccine from the governmental sector, which was also their workplace [25].

According to a study by Cowan et al. in the US, the lack of time (34%) was a common reason for not receiving the vaccine [17].

According to Wallace et al. 5–60% of the healthcare workers reported time constraints for rejection of the influenza vaccine [26].

Additionally, social support from family and colleagues plays a significant role in enhancing a person’s self-efficacy. Takayanagi et al. showed that “believing that most departmental colleagues had been vaccinated” (P < .0001) was significantly associated with compliance with influenza vaccination [24].

Among male primary care physicians, those who had received five or more influenza vaccinations in the past showed a significant positive association with self-efficacy.

Alenazi et al. found that increasing age, longer work duration in health services, being male and being a physician, are associated with significantly larger vaccination compliance (p=0.02, p=0.07, p=0.01, p=0.01) respectively [27].

The survey found no statistically significant relationship between having a history of chronic diseases and self-efficacy. This is contrary to a study by AlMarzooqi et al. where it was noted that participants with diabetes, bronchial asthma and obesity had increased acceptance of the influenza vaccine (P < .001) [25].

There is a notable difference in how physicians acknowledge risk versus their perception of risk. Most physicians agree that healthcare professionals are at the highest risk for influenza during epidemics, however, fewer believe they are at high risk. Studies have found that one main reason for not receiving seasonal influenza vaccine among healthcare professionals was that they did not recognize themselves as at risk [28,29].

According to AlMarzooqi et al. the most common reason among health care professionals for not being vaccinated was their belief of not at high risk of contracting influenza [25].

The study revealed that physicians primarily depend on official sources for information about the influenza vaccine, such as the Ministry of Public Health, WHO, and CDC. In contrast, they consult electronic and print media and flu-specific resources much less frequently for influenza information. Alame et al. reported the participants relied on various sources for vaccine recommendations, including international organizations (e.g., WHO), and the Ministry of health [30].

Most healthcare professionals (70.5%) were aware of CDC recommendations concerning influenza vaccines for healthcare professionals [25].

## Conclusion

This study highlights gaps between professional risk acknowledgment and personal risk mitigation, underscoring the need for targeted strategies to address vaccine hesitancy, improve access, and align perceptions with evidence-based practices in healthcare settings

## Data Availability

All relevant data are within the manuscript and its Supporting Information files.

## Acknowledgments

The authors of this manuscript would like to thank primary care physicians in Qatar who participated in this survey.

